# Impact of control strategies on COVID-19 pandemic and the SIR model based forecasting in Bangladesh

**DOI:** 10.1101/2020.04.19.20071415

**Authors:** Mohammad Mahmudur Rahman, Asif Ahmed, Khondoker Moazzem Hossain, Tasnima Haque, Anowar Hussain

## Abstract

**Background:** COVID-19 is transmitting worldwide drastically and infected nearly two and half million of people so far. Till date 2144 cases of COVID-19 is confirmed in Bangladesh till 18^th^ April though the stage-3/4 transmission is not validated yet.

**Methods:** To project the final infection numbers in Bangladesh we used the SIR mathematical model. Confirmed cases of infection data were obtained from Institute of Epidemiology, Disease Control and Research (IEDCR) of Bangladesh

**Results:** The confirmed cases in Bangladesh follow our SIR model prediction cases. By the end of April the predicted cases of infection will be 17450 to 21616 depending on the control strategies. Due to large population and socio-economic characteristics, we assumed 60% social distancing and lockdown can be possible. Assuming that, the predicated final size of infections will be 3782558 on the 92th day from the first infections and steadily decrease to zero infection after 193 days

**Conclusion:** To estimate the impact of social distancing we assumed eight different scenarios, the predicted results confirmed the positive impact of this type of control strategies suggesting that by strict social distancing and lockdown, COVID-19 infection can be under control and then the infection cases will steadily decrease down to zero.

## INTRODUCTION

Coronavirus disease 2019 (COVID-19) is exhibiting an unparalleled challenge before the mankind. Till date (21^st^ April 2020) there are 2.48 million confirmed cases of COVID-19 and more than170K reported deaths globally[1]. Nearly 40% of the world populations are currently under lockdown by Government or community to reduce the transmission of this extreme contagious disease. COVID-19 is the viral infectious disease caused by the SARS-CoV-2 and is transmitted by respiratory droplets and fomites with incubation period from 2 to 14 days[2].Institute of Epidemiology, Disease Control and Research (IEDCR) first reported a COVID-19 case in Bangladesh on March 8, 2020[3]. There has been a steady increase in the number of infections with 2948 cases on April 20, among which there are 2,762 active cases, 85 recovered cases and 101 deaths. To combat, Bangladesh has employed international travel bans and a gradual lockdown. However, countries like Bangladesh are at a greater risk because of large population density, inadequate infrastructure and healthcare systems to provide required support. Initially, it was thought that hot and humid weather[4,5], a large proportion of the young population, and probable immunity caused by BCG vaccinations[6] may help to keep the number of infection low. However, larger portion of these outcomes are preliminary and correlation-based, thus additional confirmation is necessary for definite conclusion[7].

The trends of the maximum of pandemics follow the rapid exponential growth during the preliminary stage and ultimately fallen down[8]. The models are based on an exponential fit for short term and long term predictions. The Susceptible-Infectious-Recovered (SIR) compartment epidemiological model[9] has been used by considering susceptible, infectious, and recovered or deceased status of individuals during pandemics. Bangladesh announced a countrywide lockdown excepts the emergency services till 25 April. There is no authentic study so far how many people maintaining social distances in Bangladesh, although the current authors’ previous study(elsewhere submitted)[10] showed that 12.03% did not maintain social distances. However, the study was conducted through online cross-sectional study where a big portion of the population was not included due to unavailability of internet. Infection numbers were predicted by assuming that, 50%, 60%, 70%, 80%, 90%, 99% and 99.50% of the Bangladeshi population maintained strict social distances. Moreover, socioeconomic conditions in a country of more than 160 million populations with high density cause considerable challenges in implementing strict social distancing. Considering this fact and to know the effects of percentages of differences of social distance, two scenarios were assumed. The study estimated the prediction of highest infection cases where 99% and 99.50% people maintained strict social distancing. Different diagnostic strategies are considered in different countries for the confirmation of COVID-19 cases. In Bangladesh, in the beginning, testing was limited to persons travelling from infected countries and person did come to their direct contact. Very recently, countrywide testing has started with the suspected persons as well as selected Pneumonia patients and symptomatic healthcare workers. As on April 20, Bangladesh has tested 26,604 samples (162/million)[1]. A number of recent studies[11] have shown that the effectiveness of coronavirus infection may vary due to the warmer weather. In addition, differential immunity of Bangladeshi people due to BCG vaccine[6] was considered. Present SIR model forecasts that, the transmissions are based as a result of stage-1 (persons with a travel history to infected areas/countries) and stage-2 (person-to-person contact). However, if the confirmed cases of infections start to surpass the predicted infection thoroughly, then the outbreak will enter a new stage, and no mathematical model explained here will be applicable. Nevertheless, as on April 20^th^, there is no strong evidence for community transmission.

High population density as well as socio-demographic characters puts Bangladesh on a high risk for stage-3 and stage-4 community transmission. Although, the social distancing and scrupulous contact tracing actions are taken by the authority of Bangladesh, still there are few limiting factors. These factors need to be measured for reaching conclusions through the present SIR model study. The missing expat populations with infections possibly will also influence the predictions but this could be a debatable issue.

The most important and common questions regarding COVID-19 is its final infection numbers and death tolls. To know the answer, a range of mathematical epidemic models viz. stochastic[12], analytical[13] and phenomenological have been used[14]. In the present study, it was attempt to estimate the final epidemic size of COVID-19 using the classic compartmental Susceptible-Infected-Recovered (SIR) model [9]. With this model, a series of predictions with different circumstances regarding impact of control strategies on COVID-19 was forecasted.

## METHODOLOGY

### Data retrieval

All the data used in this study were obtained from IEDCR[3]. Present population of Bangladesh on dated 14^th^ April 2020 was retrieved from the website Worldometers.info[15].

### SIR Model

To predict the maximum infection number, SIR Epidemic Model[9]was used. The SIR Epidemic Model is a method of modeling infectious diseases by categorizing the population based on their disease condition. This classifies susceptible, infected and recovered patients. The susceptible population means they are not affected, however, are at risk for infection. Infected persons already infected by the causative agents and are able to infect the susceptible persons. Recovered means infected persons who have either recovered from the disease or achieved stable immunity, or are otherwise detached from the population that are not able to infect susceptible population (death, quarantine etc.). The SIR model presents the increased or decreased information of an outbreak based on some initial data i.e. total given population (N), the infection rate of the infectious disease (β), the recovery rate of the disease (□), initial susceptible population (S_0_), initial infected population (I_0_) and the initial recovered population (R_0_).

This model assumes blocked populations where no one dies or born, so the population remains constant and every person is either part of S, I, or R. The general form of the model is:

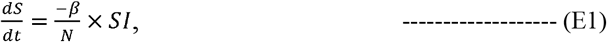

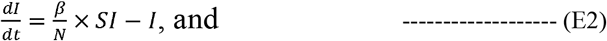

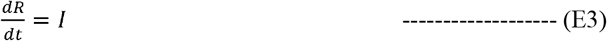

Here, β is infection rate per day, N is the total given population, and □ is the recovery rate per day. (Thus, ^<u>1</u>^ is the mean infection time). Again 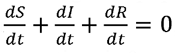, and S_0_+I_0_ +R_0_ =N indicate that the population is closed and change in numbers with respect to time is 0.

Some initial conditions viz. the initial susceptible numbers, infected numbers and recovered populations were considered to use this model. That means:

➢ S_0_ > 0 (population who are susceptible),
➢ I_0_ > 0 (at least one infected that can infect susceptible persons), and
➢ R_0_ ≥ 0 (there may be some people already recovered or died population at the start of the model, or there may be no one).

Again, S_0_+I_0_+R_0_=N and S_t_+I_t_+R_t_=N for any t. Since R_t_ can be found exclusively based on S_t_ and I_t_, considering these variables, it can be written as:

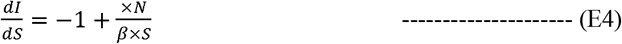

After integration,

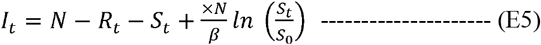

As S_t_ decreases with t increases (susceptible persons are infected but not ever added back into the susceptible numbers) and 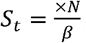, is the maximum value of I_t_ and if 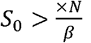 then I_t_ will raise to that highest before declining to zero. However, in case of 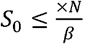 then I_t_ will decline to zero and there will be no epidemic. It can certainly be said that I_t_ must approach to zero as t→∞, and while it is observed from the model 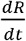 is, strictly positive and based only on I_t_. Therefore, if I_t_ was not zero, R_t_ would increase freely, which is not possible because the population is blocked.

The dynamics of the SIR mathematical model depend on the ratio:

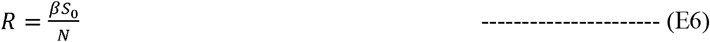

Where, R is the effective rate. R is *R*_0_ = ^*<u>β</u>*^, referred to as the basic reproduction ratio or basic reproduction number. As β is the infection rate per day, and ^<u>1</u>^ is the average infectious time (or average time an individual stays infected). Generally, if R_0_>1, then infected persons are transmitting diseases into susceptible people quicker than recovery rate, so the disease grows to be an epidemic. If R_0_<1, an epidemic does not take place.

A numerical method for solving SIR differential equations:

In a compartmental model, SIR, populations are moving from one compartment to another. This model can often be molded using recursive interaction of the form, thus we can write:

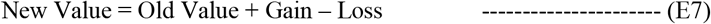

Considering these parameter, the number of population at any time who are susceptible, infected or recovered may be calculated with the following equations. These equations estimate the number of person in each state today (n), based on the number yesterday (n-1) and the rates of infections and recovery β and, respectively. The n denotes the number in one-time period and n-1 stands for the number in the prior period.

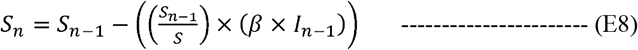

Therefore, with a time period of one day, the equation eight (E8) can be explained as the number of susceptible individual today (S_n_) equals the number of yesterday (S_n-1_), minus the fraction of people who turn into infected today (yesterday’s number of susceptible individual (S_n-1_) divided by the original susceptible number (S), multiplies their rate of infection *β* and number of individuals were infected (I_n-1_) yesterday.

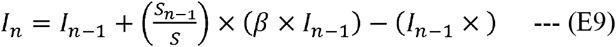

In the equation number nine (E9), the numbers of infected person today (I_n_) equals to the numbers who were infected yesterday (I_n-1_), in addition, the numbers of susceptible individual who became infected today, and subtract the numbers of recovered today who were infected yesterday.

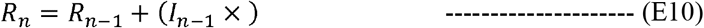

In equation number ten (E10), the number of recovered people today (R_n_) equals to the previous numbers who had recovered and the numbers who were infected yesterday and recovered today.

### Analysis

COVID-19 infection data for Bangladesh were obtained from ICEDR[3]. The total population of Bangladesh[15] was considered for this study. All the analyses were done in Microsoft Excel 2012 and SPSS 26 using the equations (E8, E9, and E10).Graphs were prepared using GraphPad Prism 8.

## RESULTS

### Current outbreak trends

According to IEDCR, on March 8, three individuals were confirmed with COVID-19 infection. Since then infection cases are gradually increasing and till date April 20 it reached to 2948 (Figure 01 a).

**Figure 01:**
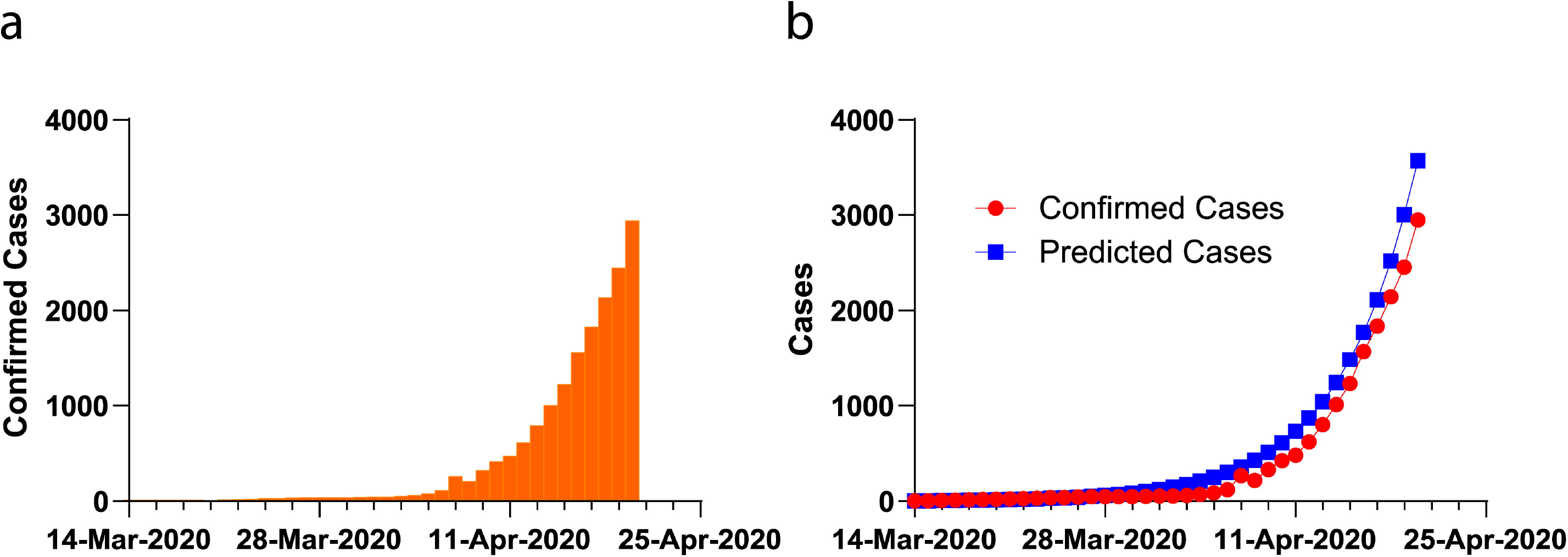
(a) Confirmed cases of infections. According to IEDCR [3], till 20th of March, the total confirmed cases were 2948 from the test of 26604 suspected cases. (b) Comparison of predicted cases and confirmed cases of infection till April 18. The confirmed cases of infection steadily followed the predicted cases obtained the SIR model analysis. The correlations are confirmed with chi square test where R2 = 0.870 and P<0.001

### Prediction Model

It was considered that, the SIR model can provide good forecast for the stage-1 and stage-2 infections as it was assumed that, there is no stage-3 transmission yet. In addition, it was estimated that, all cases to be symptomatic since estimation of asymptomatic cases in numbers would be difficult to consider. This possibly misjudges the real numbers of cases. Figure 01 (b) is showing the predicted and confirmed cases till April 20 suggesting that confirmed infection cases are following the SIR model prediction trends (R^2^ = 0.870, P<0.001). Combined prediction results according to the SIR model have been showed in Figure 02 (a).

**Figure 02:**
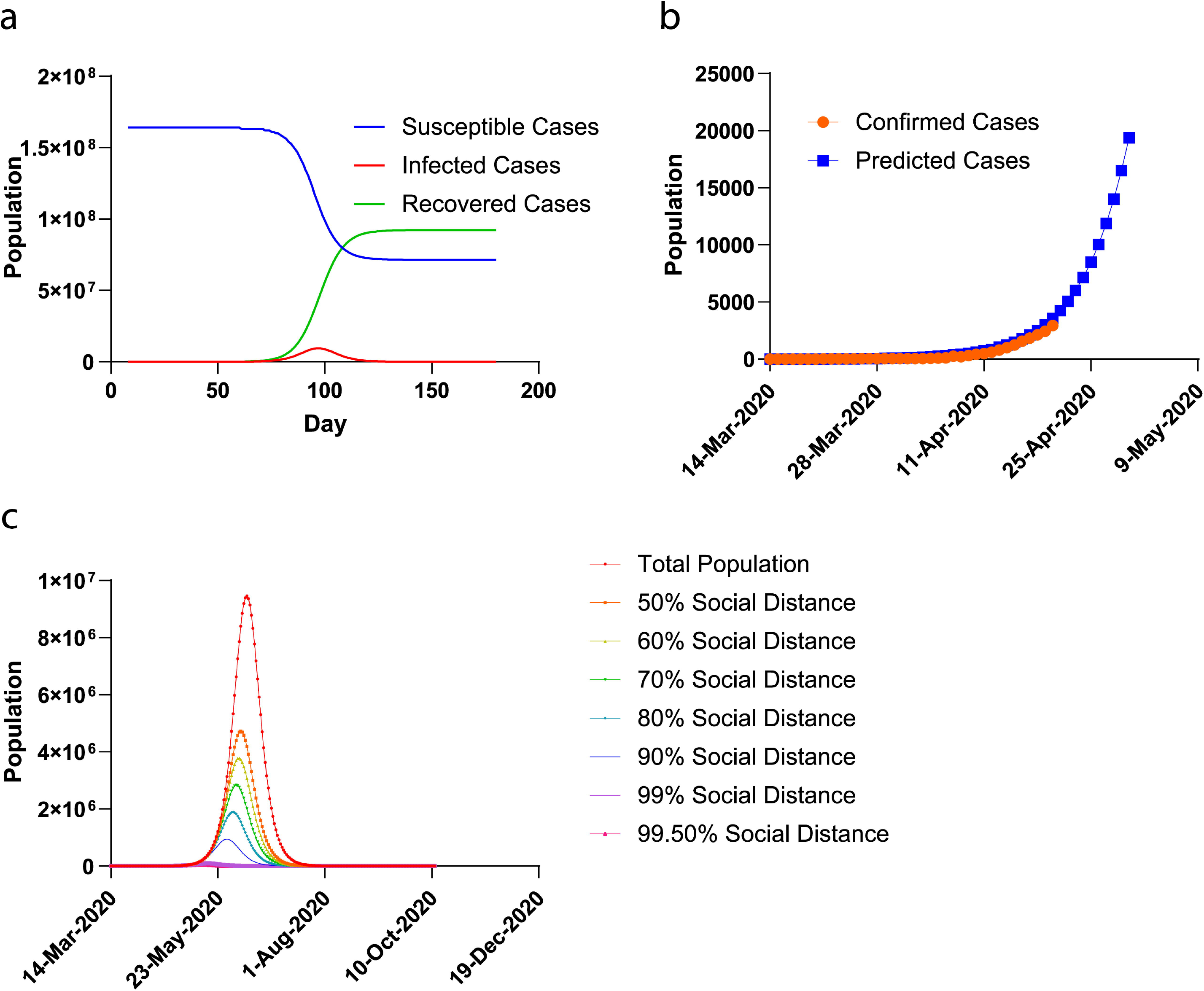
(a) The SIR Model Prediction of the COVID-19 of Bangladeshi Population with total population [15].The blue line indicates the susceptible population, green line is for recovered population and the red line indicates the infected population in SIR model analysis. Per day infection rate (0.625) was considered. (b) Prediction of infection cases at the end of April. Confirmed cases till 20th April followed the SIR model prediction. (c) Comparison of predicted infections using SIR Model regarding eight different scenarios confirming the positive effect of strict control strategies like social distancing and lockdown

The SIR model based prediction of infection curve was compared with the confirmed cases (Figure 01 b). The comparison suggests that the confirmed cases followed the predictions till April 20. The infection case numbers at the end of April was also predicted. The prediction estimated that infections will reach to 21,616 and the whole populations are susceptible. Figure 02 (b) shows the prediction of infections along with the confirmed cases by the end of April.

### Effect of social distances and lockdown

The consequences of social isolation on COVID-19 pandemic have been observed by several investigators using diverse mathematical models[2,6]. It is well-known that the effects of social distancing become evident solitary after some days from the lockdown. Since the signs of the COVID-19 characteristically take 5-6 days to appear after SARS-CoV-2 infection, Bangladesh announced lockdown pretty early (17 days first case and the number of cases were 39) compared to China (on 830 cases) and India (on 536 cases)[17]. However, considering holidays, nearly 9 millions of people moved to countrywide from the capital City Dhaka. In addition, most of expats who returned to Bangladesh from infected regions did not follow home quarantines and they scattered in different parts of the country. As a result, the exact scenarios of maintaining social distances in Bangladesh are indistinguishable. It was further assumed that, eight possible scenarios, where 100%, 50%, 40%, 30%, 20%, 10% 1% and 0.50% Bangladeshi are susceptible on the basis of current SIR model. Figure 02 (c) and Table 01 predicted that by social distancing, COVID-19 infection cases can be controlled and reduced as well as the ending of the outbreak will be rapid. The predicted infection cases have been summarized in Table 02 during the end of April, May, June, July and August. Comparing with total population with all possible scenarios suggested that COVID-19 transmission cannot be stopped now; however, it could be decreased at tolerable level by maintaining strict social distancing.

**Table 01:** The SIR Model analysis of Bangladeshi population with different scenario.

|  | In case of no intervention at all | When 50% people maintained strict social distancing | When 60% people maintained strict social distancing | When 70% people maintained strict social distancing | When 80% people maintained strict social distancing | When 90% people maintained strict social distancing | When 99% people maintained strict social distancing | When 99.50% people maintained strict social distancing |
| --- | --- | --- | --- | --- | --- | --- | --- | --- |
| **Susceptible Population** | 163689383 | 81844692 | 65475753 | 49106815 | 32737877 | 16368938 | 1636894 | 818447 |
| **Highest infection numbers** | 9459845<br>(5.779% of total BDP) | 4730216<br>(2.890% of total BDP) | 3782558<br>(2.311% of total BDP) | 2837630<br>(1.734% of total BDP) | 1891863<br>(1.156% of total BDP) | 946048<br>(0.578% of total BDP) | 94607<br>(0.058% of total BDP) | 47297<br>(0.029% of total BDP) |
| **Days need to reach highest point of infections from the first infection** | 97 | 93 | 92 | 90 | 88 | 84 | 71 | 67 |
| **Date of reaching to highest point of infection** | 11<sup>th</sup> June 2020 | 7<sup>th</sup> June 2020 | 6<sup>th</sup> June 2020 | 4<sup>th</sup> June 2020 | 2<sup>nd</sup> June 2020 | 29<sup>th</sup> May 2020 | 16<sup>th</sup> May 2020 | 12<sup>th</sup> May 2020 |
| **Days need to decrease the infection to zero from the first infection** | 203 | 196 | 193 | 189 | 185 | 177 | 150 | 142 |
| **Date of reaching to zero infection** | 25<sup>th</sup> September 2020 | 18<sup>th</sup> September 2020 | 15<sup>th</sup> September 2020 | 11<sup>th</sup> September 2020 | 7<sup>th</sup> September 2020 | 30<sup>th</sup> August 2020 | 3<sup>rd</sup> August 2020 | 26<sup>th</sup> July 2020 |

*The Table 01 shows the prediction of infection with eight different scenarios. The first column shows the predictions results if there is no intervention taken. The following columns demonstrate the predicted results if 50%, 60%, 70%, 80% 90%, 99% and 99.50% people strictly maintain social distancing and lockdown. We assumed the last two scenarios (99% and 99.50%) to show the effect of social distancing for the large population of Bangladesh*

**Table 02:** Prediction of infections till end of August 2020 with different scenario.

|  | In case of no intervention at all | When 50% people maintained strict social distancing | When 60% people maintained strict social distancing | When 70% people maintained strict social distancing | When 80% people maintained strict social distancing | When 90% people maintained strict social distancing | When 99% people maintained strict social distancing | When 99.50% people maintained strict social distancing |
| --- | --- | --- | --- | --- | --- | --- | --- | --- |
| **8<sup>th</sup> of March\*** | 3 | 3 | 3 | 3 | 3 | 3 | 3 | 3 |
| **End of March\*** | 51 | 51 | 51 | 51 | 51 | 51 | 51 | 51 |
| **End of April** | 21616 | 21591 | 21579 | 21559 | 21518 | 21397 | 19378 | 17450 |
| **End of May** | 4148873 | 3277317 | 2940692 | 2484361 | 1840632 | 916575 | 24161 | 6620 |
| **End of June** | 1271115 | 337340 | 219261 | 125586 | 57106 | 14776 | 163 | 42 |
| **End of July** | 6753 | 1735 | 1120 | 637 | 288 | 74 | 1 | 0 |
| **End of August** | 34 | 9 | 6 | 3 | 1 | 0 | 0 | 0 |

*The Table 02 presents the SIR model prediction analysis during end of April, May, June, July and August in Bangladesh. The result clearly suggests that by taking control measures like social distancing and lockdown can limit the COVID-19 infection. According to the prediction the peak infection cases will appear in May and afterwards it will go down to zero and by August 2020, Bangladesh will be free from COVID-19 infection.*
*\* denotes the confirmed cases reported by IEDCR [3].*

## DISCUSSION

In the SIR model, the number of susceptible individual today equals to the persons who were susceptible yesterday minus the numbers who become infected today. As long as the disease is spreading, the outstanding susceptible number declining each day. If two persons are infected, the chance of any person else becoming contaminated is two times higher than one individual is infected. Therefore, any estimation of the rate of transmission of the disease needs information of the infection rate per day and the number of primarily infected and originally susceptible persons. At the beginning of an epidemic, the number of persons becoming infected each day is perhaps higher than the number recovering, so, the number of infected person will increase until more person recover. The number of susceptible individual always reduce, however the number of infected and recovered person at first goes up and then turn down.

The model used in this study is data-driven, so they are as dependable as the data are. Compared to other model based studies[17] on different locations, at the beginning; the infection patterns of Bangladesh are in exponential growth stage. According to the available data, it can be predicting that the highest size of the COVID-19 outbreak using the SIR model will be nearly 94,59,845 if there is no intervention. With such a large population and existing socio-economic status, it is not possible to maintain even 80% lockdown or social distances in Bangladesh. It was assumed that, by law and enforcements and self-awareness,60% lockdown and social distances can be maintained. In accordance, the final size of COVID-19 will be 3782558 cases as per SIR model. Early strict lockdown and social distancing is the key factors for preventing COVID-19 transmissions. Studies showed that several other countries viz. UK, Germany, Italy and USA, where the stringent action was employed only after COVID-19 community transmission stage (stage-3), the outbreak became uncontainable. Additionally, different nations have different strategies as well as acquiescence levels due to several realistic considerations in enforcing the lockdown. This may have an effect on the final size of outcome. For example, the infection rates in Italy and USA are still not becoming stable after more than 30 days of lockdown. They have witnessed the uppermost percentage of death as well. Conversely, South Korea, Japan, Singapore etc. has shown significant decline by imposing lockdown[1]. In a recent predicted study[16], a reduction of infections in Australia was evident when social distancing levels went beyond 80%. Assuming the same pattern of Australia, it can be stated that, till April 30, (37 days from lockdown), very little effect of social estrangement may be observed. By this date, Bangladesh may have reported cases as many as 21579, if 40% of Bangladesh population are susceptible. This number could rise significantly if community transmission (stage-3) turns out and transmissions due to the movement of industrial workers and laborers. In addition, on May 31st, Bangladesh should observe the peak of transmission predicted 2940692, if no further strict lockdown is imposed. To reach on the predicted numbers of infection on 30^th^of May, Bangladesh should expect around 259270 patients on a single day.

A latest study by Mandal et al. [20] has revealed that social distancing can decrease cases by up to 62%.The similar prediction was made by present study model. Exponential increase is thought throughout Bangladesh to consider for the worsen-case scenario. A reduction of 90-95% can bring the condition to more convenient. In addition, if Bangladesh handles the isolation strictly, it is anticipated that the infection curve will begin flattening out soon. Qualitatively, the model shows that the epidemic is moderating, but recent data show a linear upward trend. The next few days will, therefore, be crucial regarding the direction of the epidemic. The investigative SIR model shows that, the transmission rate of COVID-19 in Bangladesh will be as high as 9.4 million. For the existing socio-economic conditions and some other practical considerations, it would not be possible to impose strictest lockdown. However, still now, the transmission rate per day and basic reproduction number for Bangladeshi people are nearly the level of global range[17]. Due to the questionable number of testing[1] compared to other countries, referencing a low transmission rate per day and therefore, the lesser basic reproduction number than the world scenario is prevailing. The SIR model assumes all the infection cases to be symptomatic, which is a limitation and could underestimate the actual cases because of an unsure number of asymptomatic cases. With this constraint, the SIR model satisfactorily predicts the cases till today (April 21). The prediction indicates that Bangladesh will enter equilibrium by the end of the first week of June with estimated total number of cases to be approximately 3782558, if no further stringent measures taken by the Government. It is projected that the effect of social distancing will be visible shortly by the end of April. However, Bangladesh is on the door to go into community transmission due to reported infringement of quarantine standard by people as well as other socio-demographic characteristics. The predictions completed using the epidemiological model in this study will be unacceptable if the transmission goes into stage-3 massively.

Therefore, it may be concluded that, SIR model is enabling us to test our understanding of the disease epidemiology especially, COVID-19 by comparing model results and observed patterns. However, the outcomes from this study are supposed to be used only for qualitative understanding and rational estimation of the nature of pandemic, but are not meaningful for any judgment making or strategy/policy change.

## Data Availability

All data in this study were obtained from publicly available sources.

## Limitation of the study

This study was conducted with available data and concluded with predictions using SIR epidemiologic model. However, the SIR model predictions will invalid if the transmission enters into stage-3. Thus, no policy making decision should be made based on this predictions except imposing of strict lockdown.

## Contributors

MMR, TH and AA conceived the study with input from KMH. MMR, TH and AA studied the equations and prepared study design. MMR led the project regarding, data collections, analysis to writing with the help of TH and AA. MMR led the solving the equations and analyzing the data with the help from TH and AA. MMR, AA and TH produced the first draft of the manuscript; KMH did put efforts regarding writings and corrections of the manuscripts. KMH and MAH added additional points in discussions. MMR, AA, KMH, TH, and MAH finalized the manuscripts after necessary corrections and obtaining suggestions from all authors. MMR, and KMH jointly supervised all the works from the beginning to the end. All authors did read and agreed unanimously to submit the manuscripts.

## Acknowledgement

Authors thank and acknowledge to all health care workers including doctors, nurses, assistants and the law enforcement authority for their diehard efforts to manage the COVID-19 pandemic conditions in Bangladesh.

